# COVID-19 Vaccine Breakthrough Infections in Veterans Health Administration

**DOI:** 10.1101/2021.09.23.21263864

**Authors:** Aditya Sharma, Gina Oda, Mark Holodniy

**Author notes:** **Correspondence** Aditya Sharma MD, US Department of Veterans Affairs, Palo Alto, CA 94304.

## Abstract

**Background:** Vaccination against severe acute respiratory syndrome coronavirus 2 (SARS-CoV-2) has been accompanied by rising concern of vaccine breakthrough due to SARS-CoV-2 variants, waning protection over time, differential vaccine effectiveness, and regional resurgence of Coronavirus 2019 (COVID-19). Characterizing the frequency and drivers of vaccine breakthrough is necessary to inform COVID-19 control efforts.

**Methods:** We performed a retrospective cohort study of vaccine breakthrough infections in fully vaccinated persons in Veterans Health Administration. We applied Cox proportional hazard models to estimate cumulative incidence, assess differences in outcomes by vaccine, and identify associations with individual characteristics as well as time-dependent geographic variation in COVID-19 incidence, proportion of delta variant, and vaccine coverage.

**Results:** Among 3,032,561 fully vaccinated persons, documented SARS-CoV-2 infection occurred in 11,197 (0.37%) and COVID-19 hospitalization occurred in 2,080 (0.07%). Compared to Ad26.COV2.S, BNT162b2 and mRNA-1273 had lower occurrence of documented SARS-CoV-2 infection (aHR 0.54, 95% confidence interval (CI) 0.51-0.58; aHR 0.36; 95% CI 0.33-0.38; respectively) and COVID-19 hospitalization (aHR 0.56, 95% CI 0.47-0.66; aHR 0.30; 0.25-0.35; respectively). Documented SARS-CoV-2 infection and COVID-19 hospitalization were associated with younger age, Hispanic or Latino ethnicity, number of comorbidities, and previous SARS-CoV-2 infection. Regional proportion of delta variant and county-level COVID-19 incidence were predictors of vaccine breakthrough events; county-level vaccine coverage was inversely associated.

**Conclusions:** Vaccine breakthrough was rare among fully vaccinated persons. mRNA-1273 and BNT162b2 were more protective against documented SARS-CoV-2 infection and COVID-19 hospitalization compared to Ad26.COV2.S. Efforts to limit COVID-19 transmission and bolster vaccine coverage would also curtail vaccine breakthrough.

## Background

Comprehensive vaccination against SARS-CoV-2 is one of the pillars in the US National Strategy for the COVID-19 Response and Pandemic Preparedness^1^. As of September 2021, US Food and Drug Administration (FDA) approved three vaccines for use in adults to prevent COVID-19 caused by SARS-CoV-2: BNT162b2 (Pfizer–BioNTech), mRNA-1273 (Moderna), and Ad26.COV2.S (Janssen)^2^. By September 10, 2021, US Centers for Disease Control and Prevention (CDC) estimated that 64.5% of US population ≥ 18 years of age and 82.2% of persons ≥ 65 years of age were fully vaccinated with one of these vaccines^3^.

Nationwide surveillance of vaccine breakthrough cases is administered by US CDC and is limited to events that result in hospitalization or death^4^. Initial reports to CDC suggested that vaccine breakthrough is rare; however, the emergence and dissemination of SARS-CoV-2 variants, in particular delta variant, has been accompanied by concern of increased vaccine breakthrough events^5,6^. Additionally, laboratory studies have demonstrated waning antibody titers in vaccinated persons, raising further concern of vaccine breakthrough over time^7^. Several studies have characterized post-vaccination COVID-19 infections^8-13^; however, studies of breakthrough infections tend to be among small cohorts, in non-generalized populations, or in areas with narrow geographic coverage. Additionally, these studies did not examine differences in post-vaccination infections by vaccine and did not account for temporal and geographic variation of the COVID-19 epidemic was well as the rapid increase in proportion of delta variant among COVID-19 cases. These limitations present challenges in developing scientifically supported policies and clinical recommendations to address vaccine breakthrough infections in the United States.

We reviewed clinical records of individuals vaccinated in the Veterans Health Administration (VHA), the largest healthcare delivery system in the United States, with the following objectives: 1. to determine the overall frequency of vaccine breakthrough infections, 2. to examine differences in vaccine breakthrough events by vaccine, and 3. to identify person-level as well as contextual characteristics associated with vaccine breakthrough.

## Methods

### Study design

Eligibility criteria included Veterans at least 18 years or older who received two doses of mRNA-1273 or BNT162b2 vaccines within the recommended timeframe listed in FDA approvals, or received Ad26.COV2.S vaccine during January 1, 2021 to August 31, 2021; residents of nursing home facilities were excluded. For each vaccinated person, we collected demographic characteristics, comorbidities, hospitalization histories, previous SARS-CoV-2 antigen and PCR test records, and type of vaccine administered from the VHA Corporate Data Warehouse. Comorbidities were summarized based on the Charlson Comorbidity Index (Supplemental Table 1).

For each patient record, county-level population density^14^, COVID-19 vaccine coverage^15^, and COVID-19 incidence^16^ were added; proportion of delta variant by Health and Human Services (HHS) region^17^ was also included. A sigmoidal curve was fitted to delta variant data to impute proportion during dates with missing data (Supplemental Figure 1). Data describing vaccine coverage, COVID-19 incidence, and proportion of delta variant were log-transformed to approximate normal distributions for use in models.

Vaccine breakthrough events were divided into two categories: 1) documented SARS-CoV-2 infection, defined as PCR or antigen positive specimen on or after 14 days from the second dose of mRNA-1273 or BNT162b2 vaccines, or after the single of the Ad26.COV2.S vaccine; and 2) COVID-19 hospitalization, defined as an admission to an acute care facility accompanied by documented SARS-CoV-2 infection either 14 days before or 3 days after date of admission and a discharge ICD 10 diagnosis code compatible with COVID-19 infection, unspecified pneumonia, or respiratory distress (Supplemental Table 2). Follow up for each person occurred until the earliest detection of an event or the end of the study period. Previous SARS-CoV-2 infection was defined as a PCR or antigen positive specimen collected at least 90 days before date of final vaccination.

### Statistical Analysis

Cox-proportional hazard models were applied to identify relationships between covariates and vaccine breakthrough outcomes. Proportion of delta variant, COVID-19 incidence, and vaccine coverage were modelled as time-dependent covariates, with new measurements for each person at 50-day intervals until date of event or censoring. Interactions between significant covariates were assessed and added in final models. Cumulative incidence of vaccine breakthrough events was estimated. P values of ≤ 0.05 were considered to indicate statistical significance. All analyses were performed in R version 4.10^18^.

This project was approved by the Stanford University Institutional Review Board (Protocol ID 47191, “Public Health Surveillance in the Department of Veterans Affairs” and written informed consent was waived.

## Results

During January 1, 2021 to August 31, 2021, 3,032,561 Veterans were fully vaccinated. Of these, documented SARS-CoV-2 infection occurred in 11,197 (0.37%) and 2,080 (0.07%) individuals had COVID-19 hospitalization in an acute care facility. The frequency of vaccine breakthrough events was bimodal, with an early peak in April 2021 followed by a period of decline until mid-June, after which events rapidly increased (Supplemental Figure 2). Vaccine breakthrough events in VHA also varied geographically, with Arkansas, Florida, and Louisiana experiencing the highest cumulative incidence of vaccine breakthrough infections by the end of the study period (Supplemental Figure 3).

The median age of vaccinated Veterans was 70 (interquartile range [IQR]: 58-76) (Table 1); the majority (2,776,045; 91.5%) were male and Non-Hispanic White (1,894,675; 62.5%). The median Charlson Comorbidity Index was 3 (IQR: 1-5). Most received mRNA-1273 (1,511,382; 49.8%) or BNT162b2 (1,293,609; 42.7%) vaccines; Ad26.COV2.S was the least commonly received (227,570; 7.5%). At the date of administration of the final dose of vaccine, the median COVID-19 incidence in the county of residence was 16 (IQR: 10-25) cases per 100,000 persons.

**Table 1.** Characteristics of vaccinated persons and cases of vaccine breakthrough infections.

|  | Vaccinated | Documented SARS-CoV-2 Infection | COVID-19 Hospitalization |
| --- | --- | --- | --- |
| Characteristic | N = 3,032,561<sup>1</sup> | N = 11,197<sup>1</sup> | N = 2,080<sup>1</sup> |
| Age (years)<sup>1</sup> | 70 (58, 76) | 68 (57, 75) | 73 (66, 79) |
| Age Group |  |  |  |
| 18-44 years old | 334,859 (11.0%) | 1,218 (10.9%) | 53 (2.5%) |
| 45-64 years old | 843,959 (27.8%) | 3,493 (31.2%) | 420 (20.2%) |
| 65+ years old | 1,853,743 (61.1%) | 6,486 (57.9%) | 1,607 (77.3%) |
| Gender |  |  |  |
| Female | 256,516 (8.5%) | 979 (8.7%) | 86 (4.1%) |
| Male | 2,776,045 (91.5%) | 10,218 (91.3%) | 1,994 (95.9%) |
| Race/Ethnicity |  |  |  |
| Hispanic or Latino | 202,327 (6.7%) | 1,039 (9.3%) | 163 (7.8%) |
| Non-Hispanic White | 1,894,675 (62.5%) | 6,659 (59.5%) | 1,273 (61.2%) |
| Non-Hispanic Black | 514,790 (17.0%) | 2,429 (21.7%) | 473 (22.7%) |
| Non-Hispanic Other | 79,693 (2.6%) | 248 (2.2%) | 38 (1.8%) |
| Non-Hispanic Unknown Race | 43,901 (1.4%) | 170 (1.5%) | 21 (1.0%) |
| Unknown Ethnicity | 297,175 (9.8%) | 652 (5.8%) | 112 (5.4%) |
| Characteristic | N = 3,032,561<sup>1</sup> | N = 11,197<sup>1</sup> | N = 2,080<sup>1</sup> |
| Myocardial Infarction | 183,744 (6.1%) | 1,221 (10.9%) | 401 (19.3%) |
| Congestive Heart Failure | 292,815 (9.7%) | 2,119 (18.9%) | 753 (36.2%) |
| Peripheral Vascular Disease | 408,764 (13.5%) | 2,212 (19.8%) | 715 (34.4%) |
| Cerebrovascular Disease | 375,151 (12.4%) | 2,101 (18.8%) | 621 (29.9%) |
| Dementia | 100,402 (3.3%) | 677 (6.0%) | 285 (13.7%) |
| Chronic Obstructive Pulmonary Disease | 809,060 (26.7%) | 4,497 (40.2%) | 1,127 (54.2%) |
| Connective Tissue Disease | 88,028 (2.9%) | 530 (4.7%) | 153 (7.4%) |
| Peptic Ulcer Disease | 95,037 (3.1%) | 623 (5.6%) | 184 (8.8%) |
| Liver Disease | 305,456 (10.1%) | 1,876 (16.8%) | 441 (21.2%) |
| Diabetes Mellitus | 974,082 (32.1%) | 4,831 (43.1%) | 1,196 (57.5%) |
| Hemiplegia | 47,322 (1.6%) | 319 (2.8%) | 108 (5.2%) |
| Moderate to Severe Chronic Kidney Disease | 412,625 (13.6%) | 2,442 (21.8%) | 789 (37.9%) |
| Solid Tumor, Leukemia, or Lymphoma | 484,311 (16.0%) | 2,487 (22.2%) | 704 (33.8%) |
| HIV | 18,955 (0.6%) | 154 (1.4%) | 31 (1.5%) |
| Charlson Comorbidity Index | 3 (1, 5) | 4 (2, 7) | 7 (4, 9) |
| SARS-Cov-2 Infection ≥ 90 Days Before Vaccination | 43,400 (1.4%) | 267 (2.4%) | 74 (3.6%) |
| Characteristic | N = 3,032,561<sup>1</sup> | N = 11,197<sup>1</sup> | N = 2,080<sup>1</sup> |
| COVID-19 Vaccine Received |  |  |  |
| Ad26.COV2.S | 227,570 (7.5%) | 1,296 (11.6%) | 201 (9.7%) |
| mRNA-1273 | 1,511,382 (49.8%) | 4,001 (35.7%) | 684 (32.9%) |
| BNT162b2 | 1,293,609 (42.7%) | 5,900 (52.7%) | 1,195 (57.5%) |
| County Population Density (persons/km<sup>2</sup>)<sup>1</sup> | 156 (45, 470) | 199 (73, 565) | 219 (74, 600) |
| County COVID-19 Incidence (cases/100,000 persons)<sup>1</sup> | 16 (10, 25) | 17 (10, 27) | 19 (12, 31) |
| County Percent Adults Fully Vaccinated (%)<sup>1</sup> | 13 (5, 24) | 10 (3, 21) | 7 (2, 16) |
| Regional Proportion of Delta Variant (%)<sup>1</sup> | 0.01 (0.00, 0.10) | 0.00 (0.00, 0.06) | 0.00 (0.00, 0.02) |

Characteristics are expressed as number (%) unless otherwise noted.
<sup>1</sup> Median, interquartile range

Compared to all vaccinated persons, those with documented SARS-CoV-2 infection had younger age (median 68, IQR 57-75 years old) had higher Charlson Comorbidity Index (median 4, IQR 2-7), were more frequently Hispanic or Latino (1,039; 9.3%) or Non-Hispanic Black (2,429; 21.7%), had higher frequency of vaccination with Ad26.COV2.S (1,296, 11.6%) or BNT162b2 (5,900; 52.7%) vaccines and lower frequency of vaccination with mRNA-1273 (4,001; 35.7%), resided in counties with higher population density (median 199, IQR 73-565 persons per km^2^) and lower vaccination coverage (10%; IQR 3-21%). Individuals with COVID-19 hospitalization were older age (median 73; IQR 66-79), were more commonly male (1,994; 95.9%), had the highest Charlson Comorbidity Index (median 7; IQR 4-9), and the lowest vaccine coverage (7%; IQR 2-16%). Regional proportion of delta variant at date of final vaccination was low (< 0.1%) across all groups. Generally, comorbidities were more prevalent in COVID-19 hospitalization compared to documented SARS-CoV-2 infection.

### Adjusted hazard ratios

Age was inversely associated with documented SARS-CoV-2 infection (aHR 0.65; 95% CI 0.63-0.66; p < 0.001) and COVID-19 hospitalization (aHR 0.87; 95% CI 0.82-0.93; p < 0.001) (Table 2). Race/Ethnicity other than Hispanic or Latino was inversely associated with all types of vaccine breakthrough events; Charlson Comorbidity Index was associated with documented SARS-CoV-2 infection (aHR 1.57; 95% CI 1.55-1.60; p < 0.001) and COVID-19 hospitalization (aHR 2.11; 95% CI 2.04-2.18, p < 0.001.); similarly, previous SARS-CoV-2 infection was associated with documented SARS-CoV-2 infection (aHR 1.46; 95% CI 1.29-1.65; p < 0.001) and COVID-19 hospitalization (aHR 2.04; 95% CI 1.61-2.59; p < 0.001). mRNA-1273 and BNT162b2 vaccines had significantly lower associations with both vaccine breakthrough events compared to Ad26.COV2.S; mRNA-1273 vaccine had the stronger protective effect. Vaccine breakthrough events were strongly associated with population density.

**Table 2.** Adjusted hazard ratios of vaccine breakthrough infection

| Characteristic | Documented SARS-CoV-2 Infection |  |  | COVID-19 Hospitalization |  |  |
| --- | --- | --- | --- | --- | --- | --- |
|  | aHR<sup>1</sup> | 95% CI<sup>2</sup> | p-value | aHR<sup>1</sup> | 95% CI<sup>2</sup> | p-value |
| Age | 0.65 | 0.63, 0.66 | <0.001 | 0.87 | 0.82, 0.93 | <0.001 |
| Gender |  |  |  |  |  |  |
| Female | — | — |  | — | — |  |
| Male | 0.98 | 0.92, 1.05 | 0.6 | 1.30 | 1.04, 1.62 | 0.023 |
| Race/Ethnicity |  |  |  |  |  |  |
| Hispanic or Latino | — | — |  | — | — |  |
| Non-Hispanic White | 0.75 | 0.70, 0.81 | <0.001 | 0.77 | 0.65, 0.92 | 0.003 |
| Non-Hispanic Black | 0.87 | 0.81, 0.93 | <0.001 | 0.94 | 0.78, 1.12 | 0.5 |
| Non-Hispanic Other | 0.63 | 0.55, 0.72 | <0.001 | 0.67 | 0.47, 0.95 | 0.026 |
| Non-Hispanic Unknown Race | 0.79 | 0.67, 0.92 | 0.004 | 0.54 | 0.34, 0.86 | 0.009 |
| Unknown Ethnicity | 0.54 | 0.49, 0.59 | <0.001 | 0.68 | 0.53, 0.87 | 0.002 |
| Charlson Comorbidity Index | 1.57 | 1.55, 1.60 | <0.001 | 2.11 | 2.04, 2.18 | <0.001 |
| SARS-Cov-2 Infection ≥ 90 Days Before Vaccination | 1.46 | 1.29, 1.65 | <0.001 | 2.04 | 1.61, 2.59 | <0.001 |
| Vaccine Received |  |  |  |  |  |  |
| Ad26.COV2.S | — | — |  | — | — |  |
|  | aHR<sup>1</sup> | 95% CI<sup>2</sup> | p-value | aHR<sup>1</sup> | 95% CI<sup>2</sup> | p-value |
| mRNA-1273 | 0.36 | 0.33, 0.38 | <0.001 | 0.27 | 0.23, 0.32 | <0.001 |
| BNT162b2 | 0.54 | 0.51, 0.58 | <0.001 | 0.51 | 0.43, 0.60 | <0.001 |
| County Population Density | 1.15 | 1.10, 1.19 | <0.001 | 1.21 | 1.12, 1.31 | <0.001 |
| County COVID-19 Incidence<sup>3</sup> | 1.99 | 1.90, 2.08 | <0.001 | 1.56 | 1.43, 1.70 | <0.001 |
| Regional Proportion of Delta Variant<sup>3</sup> | 2.01 | 1.91, 2.11 | <0.001 | 1.48 | 1.34, 1.63 | <0.001 |
| County Percent Adults Fully Vaccinated<sup>3</sup> | 0.97 | 0.95, 0.99 | 0.013 | 0.94 | 0.90, 0.99 | 0.012 |
| Regional Proportion of Delta Variant* County Percent Adults Fully Vaccinated | 1.08 | 1.05, 1.11 | <0.001 | 1.02 | 0.97, 1.08 | 0.4 |
| County COVID-19 Incidence * County Percent Adults Fully Vaccinated | 1.13 | 1.11, 1.16 | <0.001 | 1.15 | 1.11, 1.20 | <0.001 |
| County COVID-19 Incidence * Regional Proportion of Delta Variant | 0.82 | 0.77, 0.86 | <0.001 | 0.99 | 0.90, 1.09 | 0.8 |

<sup>1</sup> Adjusted hazard ratio
<sup>2</sup> Confidence interval
<sup>3</sup> Log-transformed

Time-dependent covariates were associated with vaccine breakthrough. Notably, general COVID-19 incidence at the county level was strongly associated with documented SARS-CoV-2 infection (aHR 1.99; 95% CI 1.90-2.08; p < 0.001) and COVID-19 hospitalization (aHR 1.56, 95% CI 1.43-1.70; p < 0.001). Similarly, the regional proportion of delta variant was strongly associated with documented SARS-CoV-2 infection (aHR 2.01, 95% CI 1.91-2.11, p < 0.001) and COVID-19 hospitalization (aHR 1.48; 95% CI 1.34-1.63; p < 0.001). Vaccine coverage was inversely associated with documented SARS-CoV-2 infection (aHR 0.97, 95% CI 0.95-0.99, p = 0.013) and COVID-19 hospitalization (aHR 0.94, 95% CI 0.90-0.99, p = 0.012). A significant interaction between vaccine coverage and COVID-19 incidence was identified across outcomes.

### Cumulative incidence

At 200 days, the unadjusted cumulative incidence of documented SARS-CoV-2 infection breakthrough infections was 0.84% (95% CI 0.81-0.87%) and of COVID-19 hospitalization was 0.16% (95% CI 0.15-0.18%) (Figure 1). Adjusted cumulative incidence stratified by vaccine demonstrated notable differences: 1. The vaccine breakthrough events occurred more frequently in persons who received Ad26.COV2.S, while least often in persons who received mRNA-1273; 2. Despite observed differences by vaccine received, after 200 days post-vaccination the cumulative incidence of documented SARS-CoV-2 and COVID-19 hospitalization was less than 4% and 0.5%, respectively (Figure 2).

**Figure 1.**
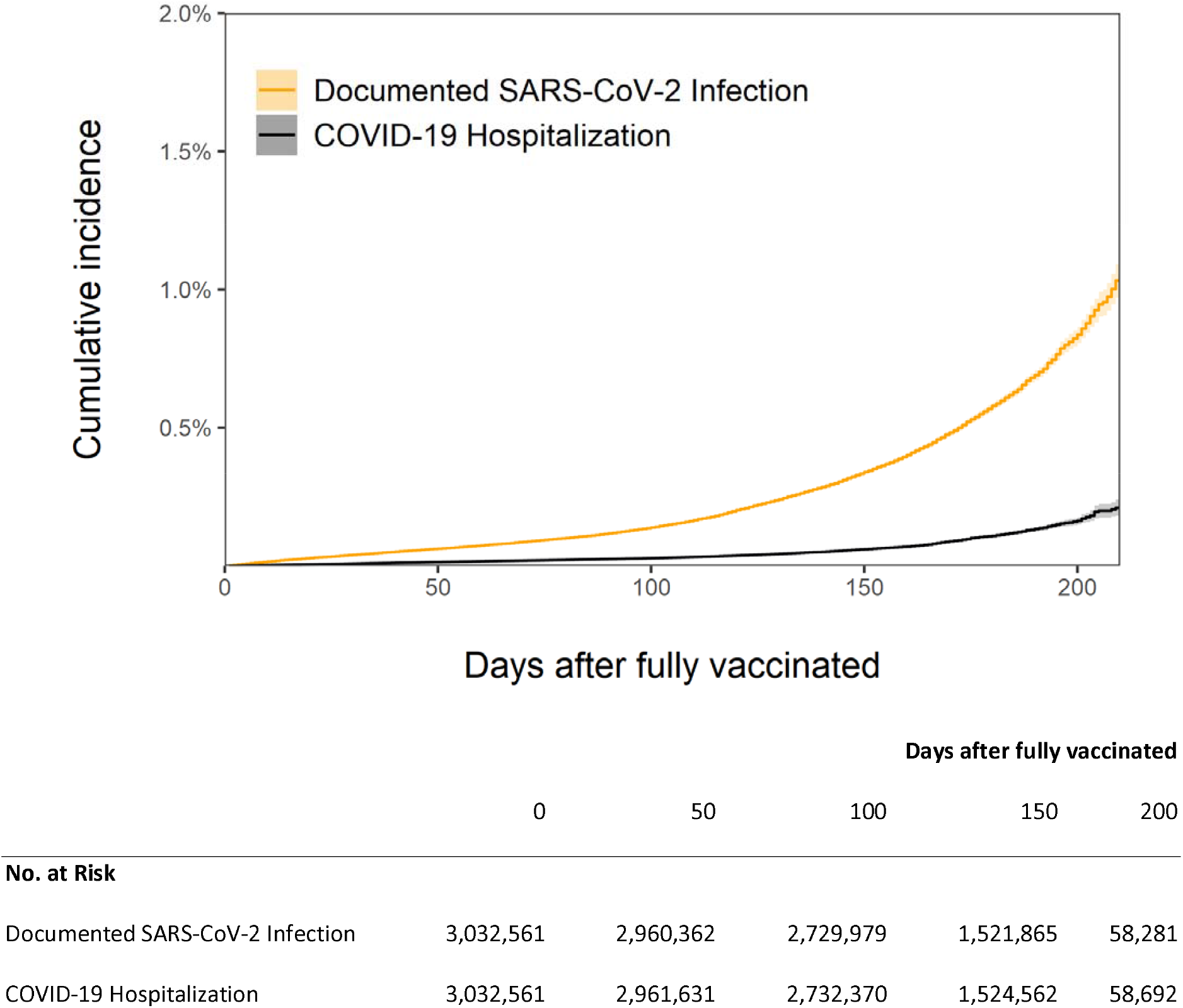
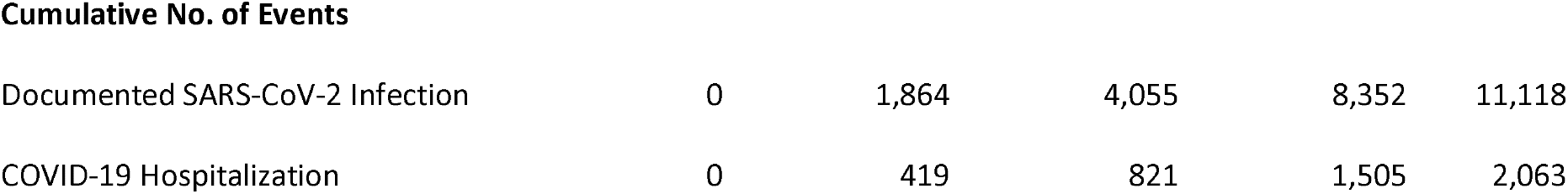
Cumulative incidence of vaccine breakthrough infections. Cumulative incidence estimates (calculated as 1 minus the survival probability) are shown for each outcome, starting from 14 days after the date of full vaccination. Shaded areas represent 95% confidence intervals. The number at risk and the cumulative number of events are also shown for each outcome at 50-day intervals.

**Figure 2.**
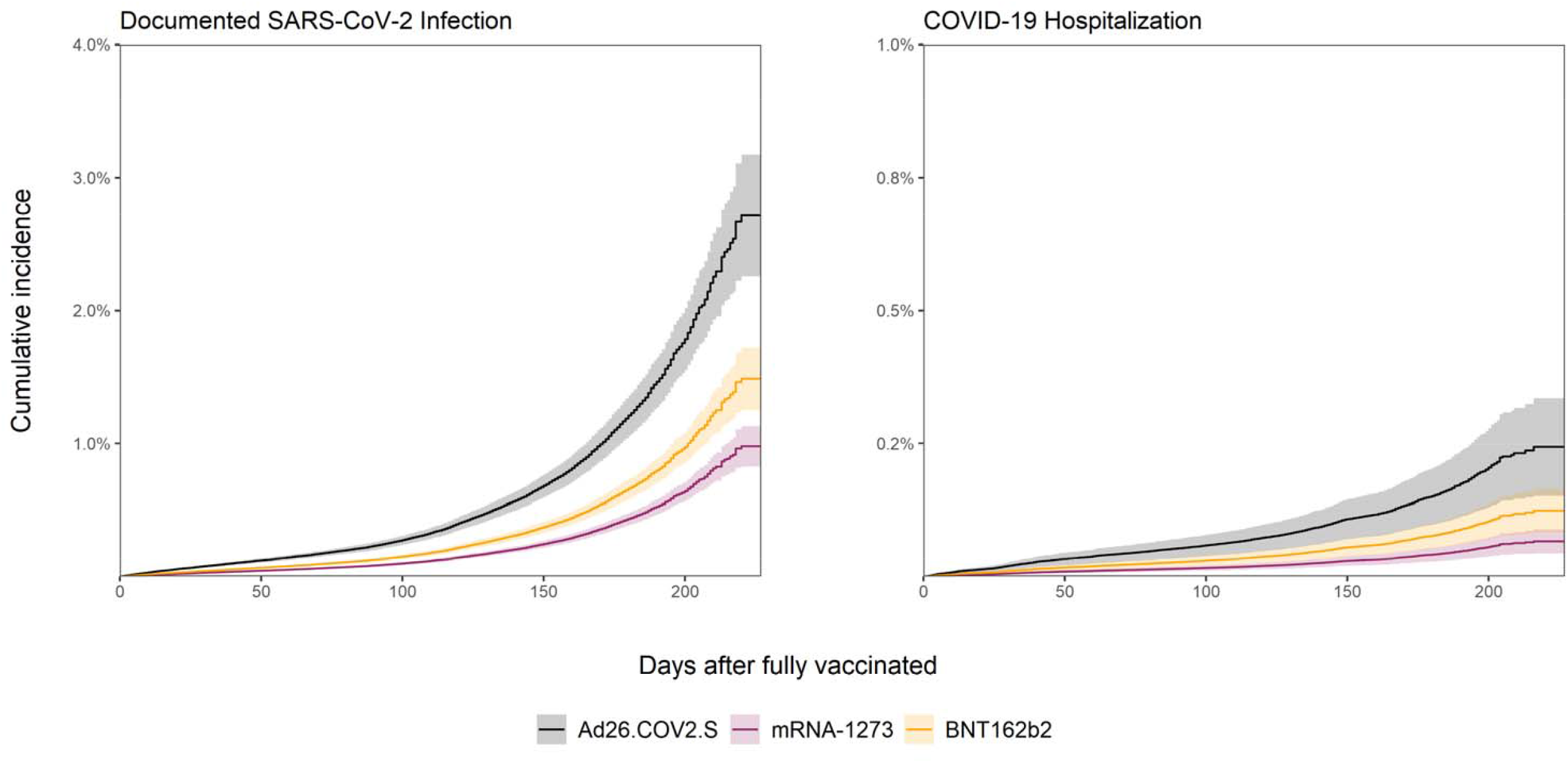
Adjusted cumulative incidence of vaccine breakthrough infections stratified by vaccine received. Cumulative incidence estimates (calculated as 1 minus the survival probability) are shown for vaccine breakthrough infections starting from 14 days after the date of full vaccination by one of three vaccines: Ad26.COV2.S, mRNA-1273, or BNT162b2; estimates are adjusted for age, sex, race/ethnicity, Charlson Comorbidity Index, previous documented SARS-CoV-2 infection, population density in county of residence, county-level COVID-19 incidence, county-level vaccine coverage and regional proportion of delta variant. Shaded areas represent 95% confidence intervals.

## Conclusions

We describe a comprehensive examination of vaccine breakthrough events in Veterans seeking clinical care across the United States. We observed significant associations between occurrence of vaccine breakthrough infection and age, comorbidities, vaccine received, documented SARS-CoV-2 infection at least 90 days before full vaccination, population density, general COVID-19 incidence in county of residence, and regional proportion of delta variant. Overall vaccine breakthrough occurrence was 0.37%, similar to an estimate described in a cohort of healthcare workers in Israel ^9^, and among fully vaccinated persons in the United Kingdom ^8^. We also observed low probability of vaccine breakthrough events across all types of vaccines – with less than 2% of vaccinated persons experiencing a vaccine breakthrough event 200 days after vaccination. Our study is the first to identify differences in vaccine breakthrough infections by type of vaccine received as well as the strong relationship between COVID-19 transmission and proportion of delta variant and the occurrence of breakthrough infections.

We observed that younger age was associated with vaccine breakthrough events. This finding might be explained by differences in behaviors among vaccinated age groups – for example, after vaccination, younger persons may be more comfortable traveling, attending gatherings, and performing other activities that could pose opportunities for SARS-CoV-2 transmission and subsequent infection^19,20^. Persons with higher number or severity of comorbidities might require more frequent encounters with clinical providers and caretakers, which could contribute to increased exposure as well as higher frequency of testing for COVID-19. Higher levels of comorbidities might also contribute to vaccine breakthrough infections due to weaker immune responses post-vaccination. The association between high population density and vaccine breakthrough infections is likely due to a combination of reduced physical distancing, increased rate of contact with COVID-19 infected persons, and higher concentrations of aerosolized COVID-19 particles compared to low population density areas^21,22^. The finding of higher occurrence of vaccine breakthrough in Hispanic or Latino persons is compatible with previous observations of increased COVID-19 in this population in part due to exposure-related factors^23^. The association between previous SARS-CoV-2 infection and vaccine breakthrough might be explained by persistence of exposures or activities related to the initial infection that continued after vaccination, or by longitudinally positive specimens in COVID-19 cases even following vaccination. Our observations regarding differences in occurrence of vaccine breakthrough by type of vaccine received are compatible with clinical trial data demonstrating higher efficacy of mRNA vaccines compared to conventional vaccines^24-26^ and also with recent observational studies comparing effectiveness of these vaccines in preventing COVID-19 hospitalizations^27^.

Evidence regarding delta variant and risk of vaccine breakthrough is mixed. In vitro studies have suggested reduced sensitivity of delta variant to neutralizing antibodies^5,6^. However, a large observational study in the United Kingdom suggests that delta variant has only modest differences in vaccine effectiveness in individuals who are fully vaccinated^28^. We observed a strong relationship between the proportion of delta variant and both forms of vaccine breakthrough; however, it is possible that this association is confounded by time-dependent contextual factors related to the rise of delta variant, such as relaxing social distancing measures or increased frequency of activities facilitating SARS-CoV-2 transmission. Additionally, we observed that vaccine breakthrough was positively associated with COVID-19 incidence and inversely associated with vaccine coverage. However, there was also a significant interaction between county-level COVID-19 incidence and vaccine coverage. Taken together, these findings suggest that the frequency of vaccine breakthrough is driven by general COVID-19 incidence after controlling for vaccine coverage; optimizing implementation of population-scale interventions to limit general COVID-19 incidence would also prevent vaccine breakthrough events. The low overall occurrence of vaccine breakthrough infections in our study suggests that boosting COVID-19 immune responses via an additional vaccine dose might not have substantial benefit in the general population^29^.

Our findings are subject to several limitations. Due to characteristics of publicly available data from the national genomic surveillance of COVID-19 variants, the geographical resolution of the proportion of delta variant was limited to the HHS regional level; it is likely that some jurisdictions would experience an earlier and more rapidly scaling proportion of delta variant than others—differential geographic dynamics in delta variant may affect the accuracy of associations between contextual variables and vaccine breakthrough events in our models. Clinical records for patients who received care in facilities external to VHA might not be available in VHA databases unless these services were ordered by VHA providers and paid for by VHA; therefore, these events would be missed in our analysis. Differences across VHA facilities in testing assays may contribute to variability in sensitivity of detection of vaccine breakthrough events; some events might have been missed or misclassified. Our study population consists of predominantly male persons and persons of older age; therefore, our results might not be generalizable to the larger US population. Given the observational nature of this study, data describing additional biomarkers, timing of exposures, symptoms, and the specific variants occurring in vaccine breakthrough events were unavailable; therefore, we were unable to assess the importance of these factors in their relationship to post-vaccination infection.

In summary, we observed that vaccine breakthrough events occurred more often in persons who received Ad26.COV2.S compared to BNT162b2 or mRNA-1273. In addition to previously observed individual-level characteristics associated with vaccine breakthrough, we observed strong associations between vaccine breakthrough events and regional COVID-19 incidence, proportion of delta variant, and vaccine coverage. Despite these findings, we observed low overall occurrence of vaccine breakthrough events more than 200 days after vaccination, suggesting a substantial protective effect against COVID-19 in persons who are fully vaccinated.

## Supporting information

STROBE checklist

## Data Availability

Due to US Department of Veterans Affairs (VA) regulations, the analytic data sets used for this study are not permitted to leave the VA firewall without a Data Use Agreement. This limitation is consistent with other studies based on VA data.

## Contributions

Conceptualization: AS, GO, MH

Data curation: AS

Formal analysis: AS

Methodology: AS, GO, MH

Project administration: AS, GO, MH

Resources: AS, GO, MH

Software: AS

Supervision: MH

Visualization: AS

Writing (original draft): AS

Writing (review & editing): AS, GO, MH

