## Supplemental appendix for "COVID-19 Vaccine Breakthrough Infections in Veterans Health Administration"

Supplementary Appendix

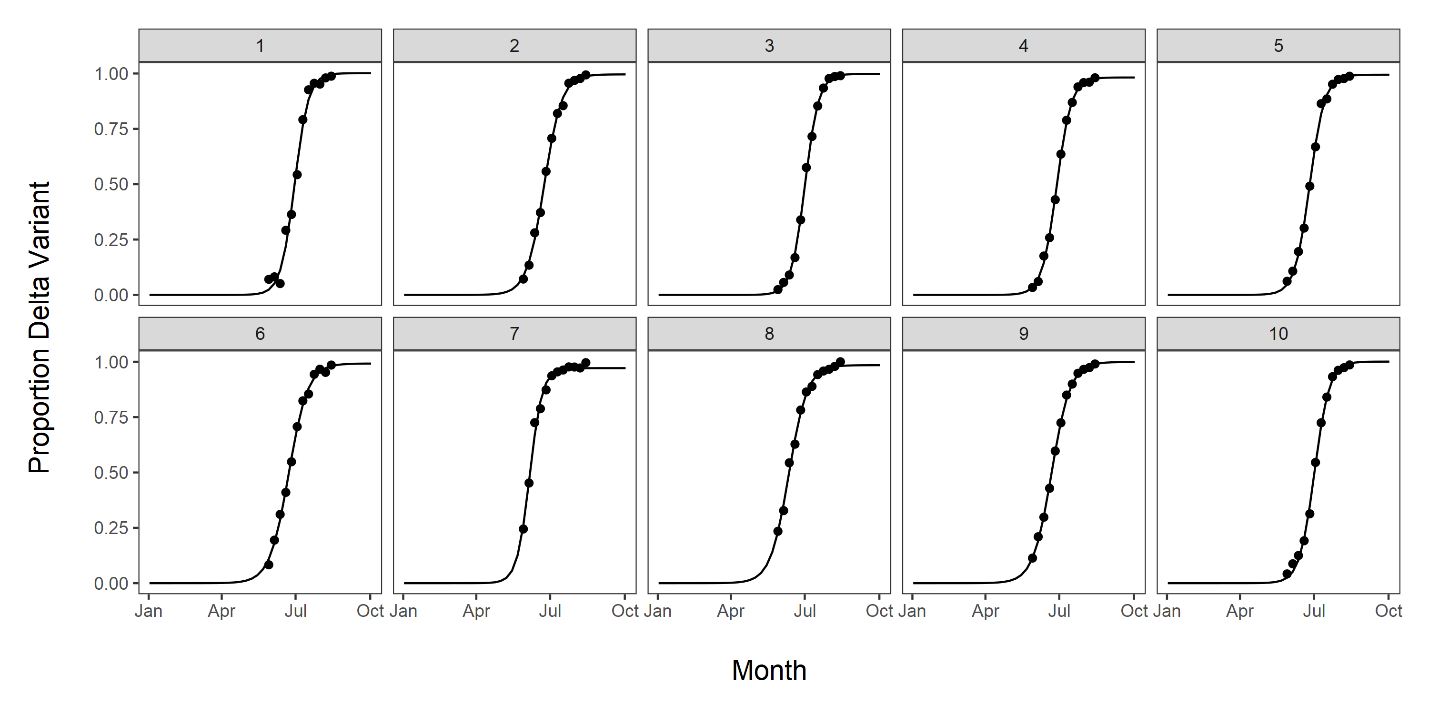

Figure 1. Fitted estimates of the proportion of delta variant by month and Health and Human Services (HHS) region. Dots represent point estimates from US Centers for Disease Control and Prevention [CDC COVID Data Tracker: Variant Proportions]. Lines depict estimates fitted to a sigmoidal curve. Labels above plot panels describe HHS region.

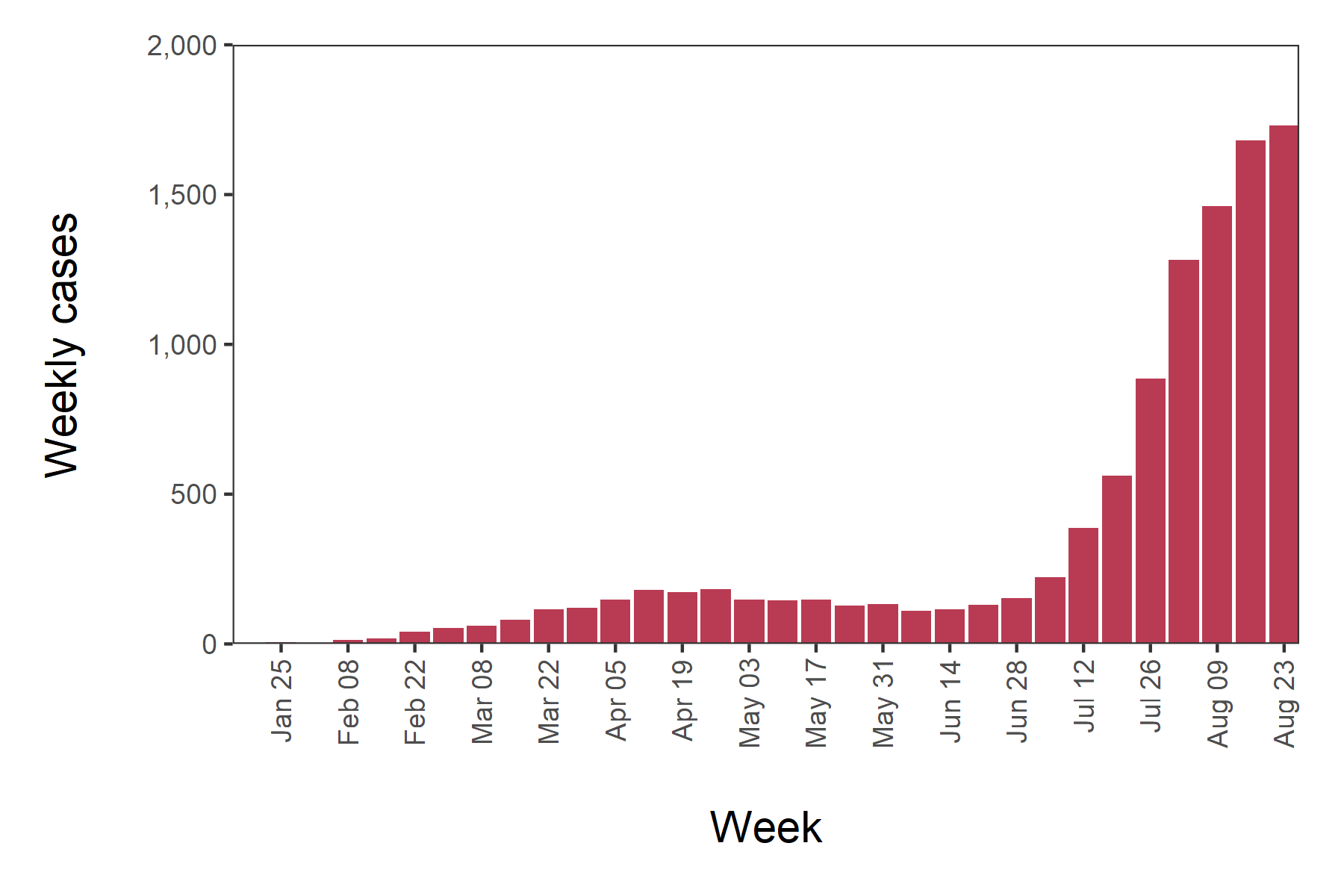

Figure 2. Total cases of all vaccine breakthrough infections in Veterans Health Administration by week. X-axis values describe starting dates for weeks during 2021. Columns represent weekly totals.

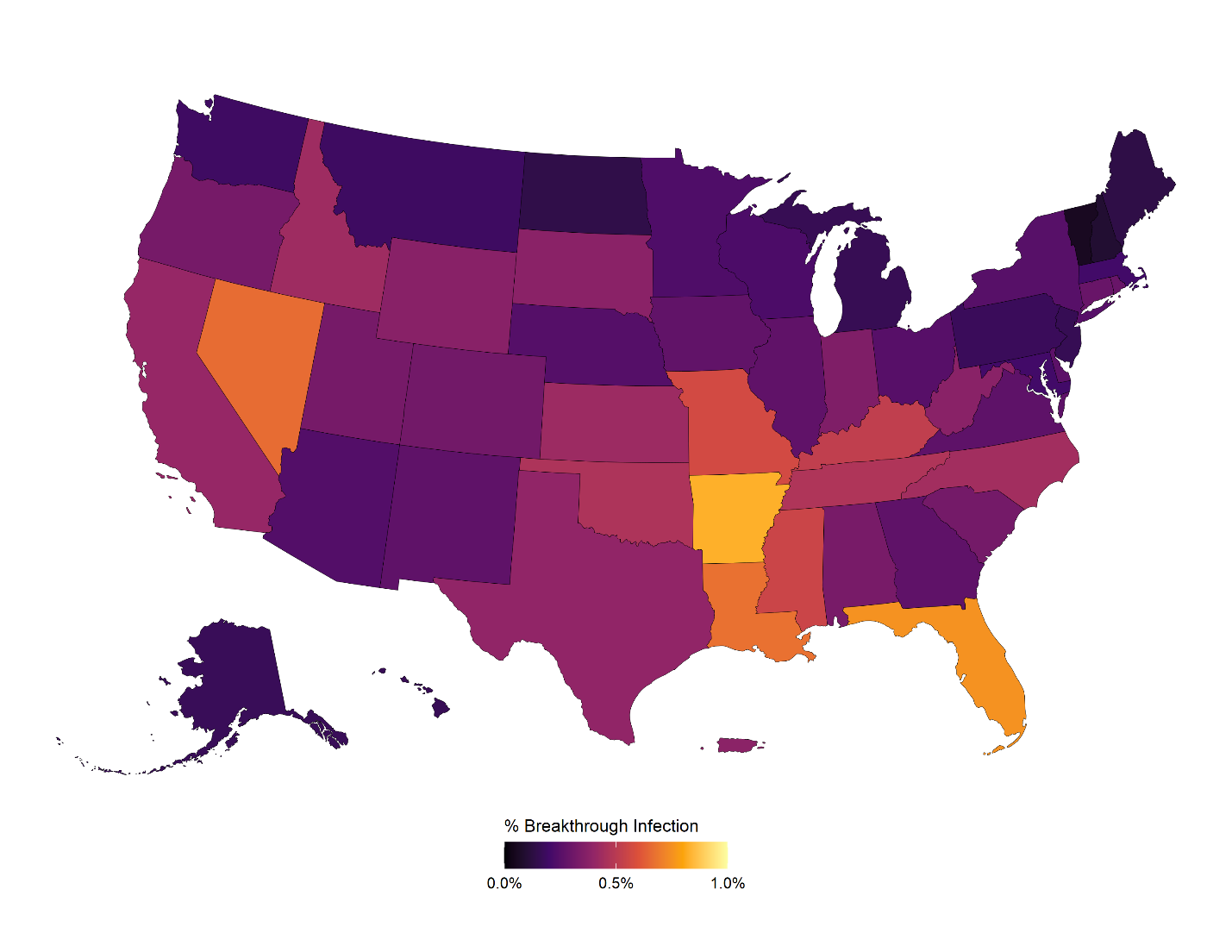

Figure 3. Cumulative incidence of vaccine breakthrough in Veterans Health Administration by state and Puerto Rico. Colors describe the percentage of all vaccinated persons with a vaccine breakthrough infection by state/territory of residence.

Supplemental Table 1. ICD-10 codes used for comorbidities based on Charlson Comorbidity Index.

| Comorbidity | ICD-10 Code |
| --- | --- |
| Myocardial infarction | I21.x, I22.x, I25.2 |
| Congestive heart failure | I09.9, I11.0, I13.0, I13.2, I25.5, I42.0, I42.5–I42.9, I43.x, I50.x, P29.0 |
| Peripheral vascular disease | I70.x, I71.x, I73.1, I73.8, I73.9, I77.1, I79.0, I79.2, K55.1, K55.8, K55.9, Z95.8, Z95.9 |
| Cerebrovascular disease | G45.x, G46.x, H34.0, I60.x–I69.x |
| Dementia | F00.x–F03.x, F05.1, G30.x, G31.1 |
| Chronic pulmonary disease | I27.8, I27.9, J40.x–J47.x, J60.x–J67.x, J68.4, J70.1, J70.3 |
| Rheumatic disease | M05.x, M06.x, M31.5, M32.x–M34.x, M35.1, M35.3, M36.0 |
| Peptic ulcer disease | K25.x–K28.x |
| Mild liver disease | B18.x, K70.0–K70.3, K70.9, K71.3–K71.5, K71.7, K73.x, K74.x, K76.0, K76.2–K76.4, K76.8, K76.9, Z94.4 |
| Diabetes without chronic complication | E10.0, E10.1, E10.6, E10.8, E10.9, E11.0, E11.1, E11.6, E11.8, E11.9, E12.0, E12.1, E12.6, E12.8, E12.9, E13.0, E13.1, E13.6, E13.8, E13.9, E14.0, E14.1, E14.6, E14.8, E14.9 |
| Diabetes with chronic complication | E10.2–E10.5, E10.7, E11.2–E11.5, E11.7, E12.2–E12.5, E12.7, E13.2–E13.5, E13.7, E14.2–E14.5, E14.7 |
| Hemiplegia or paraplegia | G04.1, G11.4, G80.1, G80.2, G81.x, G82.x, G83.0–G83.4, G83.9 |
| Renal disease | I12.0, I13.1, N03.2–N03.7, N05.2–N05.7, N18.x, N19.x, N25.0, Z49.0–Z49.2, Z94.0, Z99.2 |
| Any malignancy, including lymphoma and leukemia, except malignant neoplasm of skin | C00.x–C26.x, C30.x–C34.x, C37.x–C41.x, C43.x, C45.x–C58.x, C60.x–C76.x, C81.x–C85.x, C88.x, C90.x–C97.x |
| Moderate or severe liver disease | I85.0, I85.9, I86.4, I98.2, K70.4, K71.1, K72.1, K72.9, K76.5, K76.6, K76.7 |
| Metastatic solid tumor | C77.x–C80.x |
| AIDS/HIV | B20.x–B22.x, B24.x |

Supplemental Table 2. ICD-10 codes used for classifying hospitalizations related to COVID-19.

| ICD-10-CM Diagnosis Code | Description |
| --- | --- |
| U07.1 | COVID-19 |
| J12.82 | Pneumonia due to coronavirus disease 2019 |
| J18.9 | Pneumonia, unspecified organism |
| J96.01 | Acute respiratory failure with hypoxia |
| R06.00 | Dyspnea, not otherwise specified |
| R06.02 | Shortness of breath |
